# A meta-analysis approach to gene regulatory network inference identifies key regulators of cardiovascular diseases

**DOI:** 10.1101/2024.03.04.24303755

**Authors:** G. Pepe, R. Appierdo, G. Ausiello, M. Helmer-Citterich, PF. Gherardini

## Abstract

Cardiovascular diseases (CVDs) represent a major concern for global health whose mechanistic understanding is complicated by a complex interplay between genetic predisposition and environmental factors.

Specifically, heart failure (HF), encompassing dilated cardiomyopathy (DC), ischemic cardiomyopathy (ICM), and hypertrophic cardiomyopathy (HCM), is a topic of substantial interest in basic and clinical research. Here we used a Partial Correlation Coefficient-based algorithm (PCC) within the context of a meta-analysis framework, to construct a Gene Regulatory Network (GRN) that identifies key regulators whose activity is perturbed in Heart Failure. By integrating data from multiple independent studies, our approach unveiled crucial regulatory associations between transcription factors (TFs) and structural genes, emphasizing their pivotal roles in regulating metabolic pathways, such as fatty acid metabolism, oxidative stress response, epithelial-to-mesenchymal transition, and coagulation. In addition to known associations, our analysis also identified novel regulators, including the identification of TFs FPM315 and MOVO-B, which are implicated in dilated cardiomyopathies, and TEAD1 and TEAD2 in both dilated and ischemic cardiomyopathies. Moreover, we uncovered alterations in adipogenesis and oxidative phosphorylation pathways in hypertrophic cardiomyopathy, and discovered a role for IL2 STAT5 signaling in heart failure.

Our findings underscore the importance of TFs activity in the initiation and progression of cardiac disease, highlighting their potential as pharmacological targets.

## Introduction

Cardiovascular diseases (CVDs) are a leading cause of morbidity and mortality worldwide ^1,2^. They encompass a range of disorders involving the heart and blood vessels, including conditions such as atherosclerosis, hypertension, myocardial infarction, and cardiac arrhythmias. Among these conditions, heart failure (HF) is of major concern ^1^.

HF is a medical condition characterized by the heart’s inability to pump blood sufficiently to meet the demands of the body ^3^. HF can be caused by multiple factors, including: i) damage to the heart muscle following a heart attack; ii) uncontrolled high blood pressure; III) chronic heart disease. Symptoms of HF may include fatigue, difficulty breathing (dyspnea), and fluid buildup in the body (fluid retention) ^4^.

Multiple manifestations of HF have been described. Among these, dilated cardiomyopathy (DC) is characterized by dilation and weakening of the cardiac muscle ^5^. This condition can be idiopathic or caused by factors such as hypertension, alcohol abuse, or viral infection ^6^. On the other hand, ischemic cardiomyopathy (ICM) is a form of HF associated with inadequate blood flow to the cardiac muscle, often due to atherosclerosis of the coronary arteries ^7^. Finally, hypertrophic cardiomyopathy (HCM) is characterized by excessive thickening of the cardiac muscle, making it more difficult for the heart to pump blood ^8^.

Cardiac diseases are multifactorial disorders, heavily dependent on both genetics and environmental factors, such as individual smoking and eating habits. Recent transcriptomics studies report that a strong connection exists between such factors and the molecular mechanisms underlying cardiovascular pathologies, including HF ^9–13^. Of note, researchers showed that people’s overall lifestyles heavily affect the activity of transcription factors genes ^14,15^. Therefore, the study of gene regulatory networks will enable the discovery of fundamental mechanisms in the pathophysiology of cardiovascular diseases, and will also lead to the identification of specific transcription factors and microRNAs ^16–18^ that play a key regulatory role in health and diseases.

To this end here we employ a meta-analysis approach to reconstruct gene regulatory networks in heart failure, ischemic, dilated and hypertrophic cardiomyopathies. We leverage multiple publicly available gene expression datasets to extract different layers of regulatory information, including miRNAs, transcription factors and pathways activity. The integration of multiple datasets in a meta-analysis framework mitigates dataset-specific biases and batch effects and leads to the identification of the most robust signals in the data.

## Results and Discussion

To conduct a comprehensive and unbiased investigation of the molecular changes underlying cardiac diseases, we systematically searched the Gene Expression Omnibus (GEO) database (https://www.ncbi.nlm.nih.gov/geo/) to retrieve all the publicly available human heart disease-related bulk gene expression datasets which also incorporate microRNA expression profiles ^19^.

We collected six studies, each provided with samples from both healthy individuals and patients with different heart conditions, including heart failure, ischemic cardiomyopathy, dilated cardiomyopathy, and hypertrophic cardiomyopathy. The description of the cohorts is provided in Table 1.

**Table 1:** Summary of study cohorts.

| **GEO** | **Disease** | **# of diseased samples** | **# of healthy samples** |
| --- | --- | --- | --- |
| GSE55296<sup>20</sup> | Dilated Cardiomyopathy | 13 | 10 |
| GSE116250<sup>21</sup> | Dilated Cardiomyopathy | 37 | 14 |
| GSE133054<sup>22</sup> | Hypertrophic Cardiomyopathy | 8 | 8 |
| GSE135055<sup>23</sup> | Heart Failure | 21 | 9 |
| GSE133054<sup>22</sup> | Heart Failure | 7 | 8 |
| GSE48166 | Ischemic Cardiomyopathy | 15 | 15 |
| GSE55296<sup>20</sup> | Ischemic Cardiomyopathy | 13 | 10 |
| GSE116250<sup>21</sup> | Ischemic Cardiomyopathy | 13 | 14 |

The workflow adopted in this project is summarized in Figure 1. Initially, the gene expression profiles of individual patients were obtained. Then, differential expression between patients and healthy controls was computed for each gene. Raw gene expression values were then used to calculate higher-level, more interpretable, features such as pathways and transcription factors activity scores and their difference in patients vs controls. Our focus then shifted towards the identification of co-expressed gene pairs within the dysregulated pathways. This step involved identifying genes that exhibited coordinated expression patterns, shedding light on potential functional relationships and interactions within these molecular networks. Finally, we employed a correlation analysis within dysregulated pathways to infer specific gene regulatory networks and identify master regulators responsible for re-wiring of gene expression programs in each disease.

**Fig.1.**
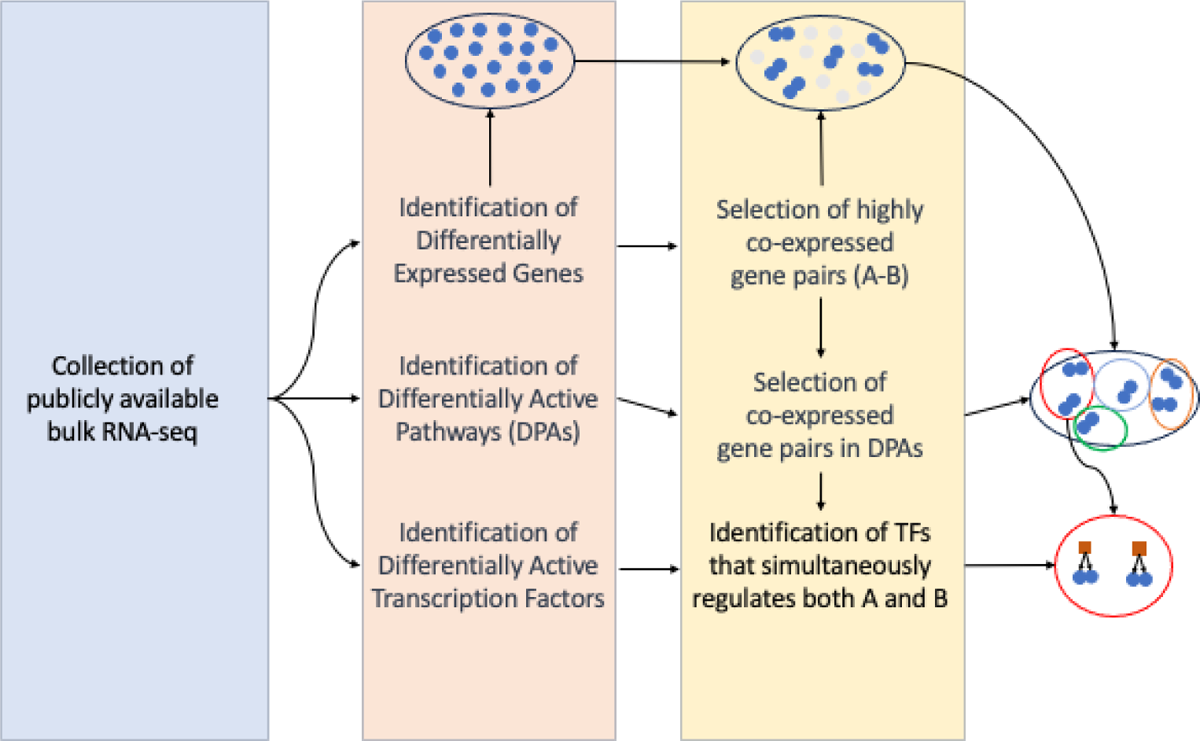
Schematic overview of the entire analysis

### Transcriptome changes in patients with cardiac pathologies

We performed differential gene expression analysis to assess transcriptomic differences in patients affected by heart failure, dilated cardiomyopathy, ischemic cardiomyopathy, and hypertrophic cardiomyopathy with respect to healthy controls. For each pathology, an integrated multi-cohort analysis was performed, which resulted in the identification of hundreds of genes that were significantly differentially expressed in disease-affected individuals compared to control individuals ( FDR <= 0.05 and an |Effect Size| >= 1). Overall, this analysis shows that only 35 genes are dysregulated in all four pathologies (Fig. 2b). The list of differentially expressed genes identified in each cardiac pathology is provided in Supplementary S1. A total of 642 differentially expressed genes (145 overexpressed and 497 underexpressed) were identified from the multi-cohort analysis performed on heart failure. Of the 145 upregulated genes, 143 were protein-coding genes and were found to be enriched in biological processes such as small molecular metabolic processes, circulatory system processes, and vascular processes in the circulatory system (Fig 2a). Additionally, these genes exhibit an enrichment of motifs recognized by transcription factors and methyltransferases (Supplementary S1), including a transcription factor already established as a prognostic biomarker for heart failure, ZNF300 ^24^. Out of the total, 2 genes were long intronic non-coding RNAs of unknown function (Fig. 2c). However, considering that in recent years several studies have linked the role of long non-coding RNAs to heart diseases ^25–28^, these long intronic non-coding genes are worthy of further investigation.

**Fig. 2.**
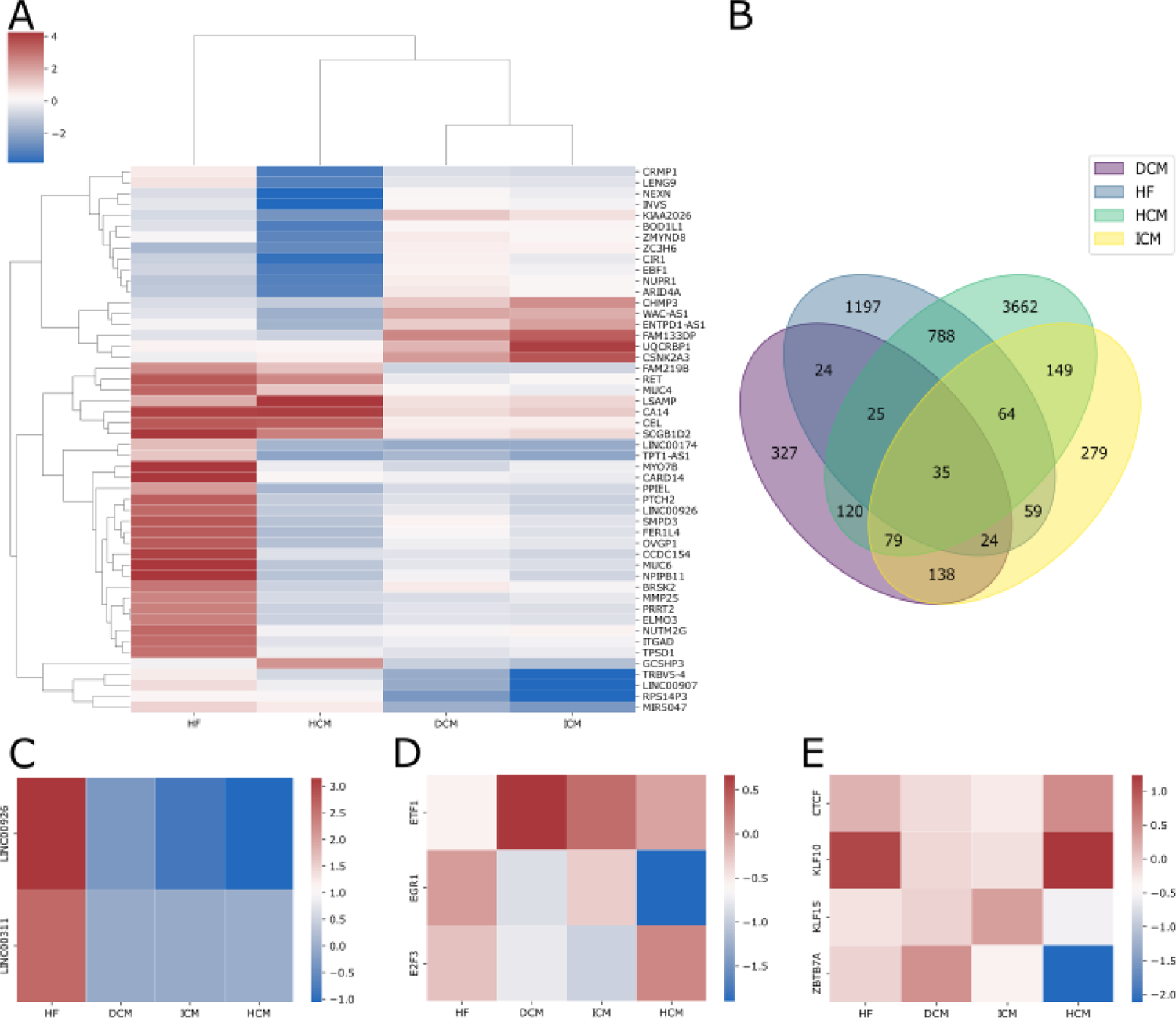
(A) The heatmap shows the pooled effect sizes resulting from the meta-analysis of genes integrated across all studies for each pathology;(B) The Venn diagram depicts the intersection of differentially expressed genes across heart failure, dilated cardiomyopathy, ischemic cardiomyopathy, and hypertrophic cardiomyopathy; (C) Effect sizes of upregulated long non-coding RNAs in heart failure patients across all four pathologies; (D) Effect sizes of transcription factors recognizing enriched motifs in dysregulated genes in dilated cardiomyopathy; (E) Effect sizes of transcription factors recognizing enriched motifs in dysregulated genes in ischemic cardiomyopathy.

On the other hand, the downregulated genes were found to be enriched in biological processes such as anatomical structure development and supramolecular fiber organization, as well as in molecular functions like extracellular matrix structural constituent, glycosaminoglycan binding, and cytoskeletal protein binding (Fig 2a). Additionally, significant enrichment was observed in the extracellular matrix organization pathway (Supplementary S1), which has been widely associated with heart failure ^29–31^.

Similarly, we conducted a multi-cohort analysis on patients with dilated cardiomyopathy, resulting in 331 differentially expressed genes (100 upregulated and 231 downregulated). Consistent with the literature, the downregulated genes were found to be enriched in biological processes such as glycoprotein metabolism, extracellular structure, or matrix organization (Fig 2a) ^32–35^. Additionally, the regulatory regions of these genes exhibit an enrichment in motifs recognized by transcription factors which have previously been observed to be involved in dilated cardiomyopathy (Fig. 2d, such as E2F3, EGR-1 (less expressed in patients) and ETF (more expressed)) ^36–38^. Furthermore, these genes display an enrichment, in their regulatory regions, for a motif recognized by the transcription factor MOVO-B (Supplementary S1), that is known to be involved in angiogenesis and heart formation ^39^. To the best of our knowledge, its role in cardiac disorders has not been previously reported ^39^. This suggests that further studies are needed to understand its potential involvement in dilated cardiomyopathy. In accordance with literature evidence, the upregulated genes are enriched in genes involved in the downregulation of the ERRBB2:ERBB3 signaling pathway ^40–42^. Considering that ERBB2 signaling regulates cardiac functionality, this pathway has been suggested to be essential for the prevention of dilated cardiomyopathy ^43^. Lastly, these genes exhibit enrichment for a promoter motif recognized by the transcription factor FPM315 (Supplementary S1), which is expressed in various tissues, including the heart, but that has not been associated with any cardiac pathology.

The multi-cohort analysis conducted on ischemic heart disease highlighted 260 differentially expressed genes, 29 upregulated and 231 downregulated. Of the 231 downregulated genes, 205 are protein-coding genes and were found to be enriched in molecular functions encompassing signaling, cell communication, and cell junction organization (Fig 2a). Additionally, the regulatory regions of these genes exhibit an enrichment in motifs recognized by transcription factors (Supplementary S1), known to be associated with ischemic cardiomyopathy, such as CTCF, KLF10, KLF15, and ZBTB7A ^44–47^. However, in our results, the expression of these transcription factors does not appear to be dysregulated in patients (Fig. 2e).

The upregulated genes were enriched in molecular functions comprising oxidoreductase activity and biological processes like the regulation of heart rate, actin-mediated cell contraction, and cardiac chamber morphogenesis (Fig. 2a, Supplementary S1). Also in this case several transcription factors appear to regulate pathogenic processes and deserve further study.

Finally, the analysis of the only hypertrophic cardiomyopathy dataset resulted in the identification of 303 differentially expressed genes (62 upregulated and 241 downregulated). The downregulated genes were enriched in biological processes such as metabolism regulation, developmental processes, and molecular functions like histone binding (Fig 2a, Supplementary S1). Among the promoters of overexpressed genes, we identified enrichment in a motif recognized by the transcription factor ZBRK1. This transcription factor is expressed in various tissues, including the heart and it has been associated with different forms of tumors, including breast cancer, hepatocellular carcinomas, and colon cancer ^48,49^. To the best of our knowledge, its involvement in heart disease has not previously been reported.

The multi-cohort analysis of cardiac pathologies showed that, in terms of differential expression profiles, ischemic and dilated cardiomyopathies are most similar to each other (Fig 2a). This is in line with previous reports that describe ischemic cardiomyopathy as a subtype of dilated cardiomyopathy ^50^. Furthermore, hypertrophic cardiomyopathy appears to be more similar to dilated cardiomyopathy. The literature corroborates this finding as well as it is known that hypertrophic cardiomyopathy, which is characterized by the thickening of the heart muscle, can sometimes evolve into dilated cardiomyopathy, which results in the enlargement and weakening of the heart chambers ^51^.

In order to get a more interpretable overview of the transcriptomic differences across the different diseases under study we next turned to the analysis of pathway activity scores.

### Alterations in pathway activity profiles

For each cardiac pathology under investigation, we conducted a multi-cohort analysis to highlight differential pathways activity between healthy and disease-affected individuals (Supplementary S2). Figure 3 shows the results for each of the four pathologies under study. In this case as well, the overall pathway activity of dilated cardiomyopathy appears to be the most similar to the one of ischemic cardiomyopathy, confirming our previous observations. The differential pathway activity provides a more interpretable view of the mechanisms involved in each pathology, and allows us to highlight differences and similarities among them. Overall, this analysis identified six pathways that are dysregulated in all four pathologies, as well as others that are specific to each disease (Fig. 3b).

**Fig. 3.**
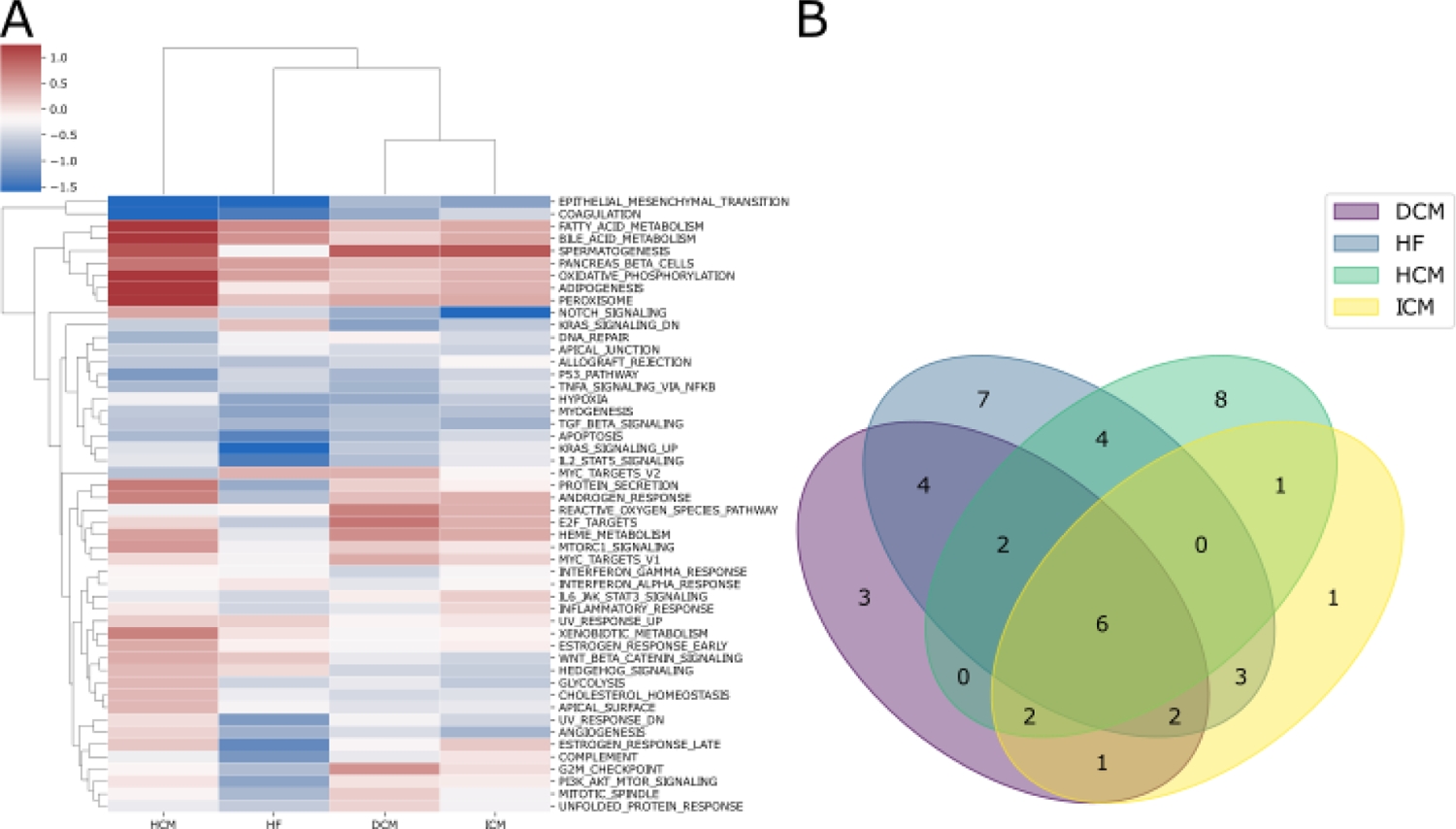
(A) The heatmap shows the pooled effect sizes resulting from the meta-analysis of pathway gene sets computed by GSVA and then integrated across all studies for each pathology using the random-effects model. This analysis further confirms the similarity between DC and ICM, now at the level of pathway activity.; (B) The Venn diagram illustrates the intersection of differentially activated pathways across heart failure, dilated cardiomyopathy, ischemic cardiomyopathy, and hypertrophic cardiomyopathy.

The epithelial-to-mesenchymal transition (EMT) and coagulation pathways exhibit a similar trend with varying degrees of dysregulation in all four pathologies. Numerous signaling pathways play a significant role in regulating EMT during cardiac development ^52,53^. The NOTCH pathway is also crucial for EMT, although it is not necessary for the initial formation of extracellular matrix swellings ^54^. Our analysis shows a downregulation of this pathway in patients with heart failure, ischemic cardiomyopathy, and dilated cardiomyopathy (Fig 3). In summary, signaling pathways involving bone morphogenetic proteins (BMPs) and TGF-β ligands and receptors, modulated by the Hippo pathway ^55^, induce the expression of Snai1, Snai2, and Twist in endocardial cells. These genes encode archetypal transcription factors that regulate EMT.

Similar to the NOTCH pathway, our analysis reveals that the TGF-β pathway is downregulated to varying degrees in the four analyzed cardiac pathologies (Fig 3).

Due to dysregulation of these signaling pathways, endocardial cells within the cushions undergo EMT and transition into a fibroblastic fate. Similar to fibroblasts found in other connective tissues, valvular fibroblasts undergo a maturation process akin to the formation of bone, cartilage, and tendons. The transcription factor SOX9, induced by BMPs, serves as a central regulator of ECM gene expression networks ^56^.

While heart failure and the three different cardiomyopathies share a common set of dysregulated pathways, there are also disease-specific trends. As reported in the literature, we observed an upregulation in pathways related to fatty acid metabolism, oxidative phosphorylation, and adipogenesis in hypertrophic cardiomyopathy (Fig 3) ^57,58^. These pathways do not appear to be upregulated in the other three pathologies, highlighting differences in metabolic reprogramming among cardiomyopathies.

Our results indicate that the K-Ras pathway is downregulated in patients when compared to healthy individuals across all four pathologies. Notably, this downregulation is more pronounced in heart failure (Fig 3a). While only a limited number of studies have explored K-Ras in cardiac research, its association with cardiac cell proliferation is evident ^59^. The identification of pathways known to be associated with cardiac pathologies validates our results, which also includes pathways, such as IL2 and STAT5 signaling or peroxisome, that, to the best of our knowledge, have not been previously implicated in cardiac disease. In order to identify key regulators responsible for the observed differences in pathway activities, we next analyzed the role of microRNAs, and the activity of Transcription Factors in these samples.

### Differentially expressed microRNAs

We performed a multi-cohort analysis to identify differentially expressed miRNAs between disease-affected and control individuals (Supplementary S3). This analysis revealed a contrasting trend in the abundance of differentially expressed miRNAs across the four diseases. No common dysregulated miRNAs were identified across the four pathologies and significant differences were observed in the number of dysregulated miRNAs in each disease. Specifically, only 5 and 22 miRNAs were found to be differentially expressed in DCM and IICM respectively (Fig 4b).

**Fig 4.**
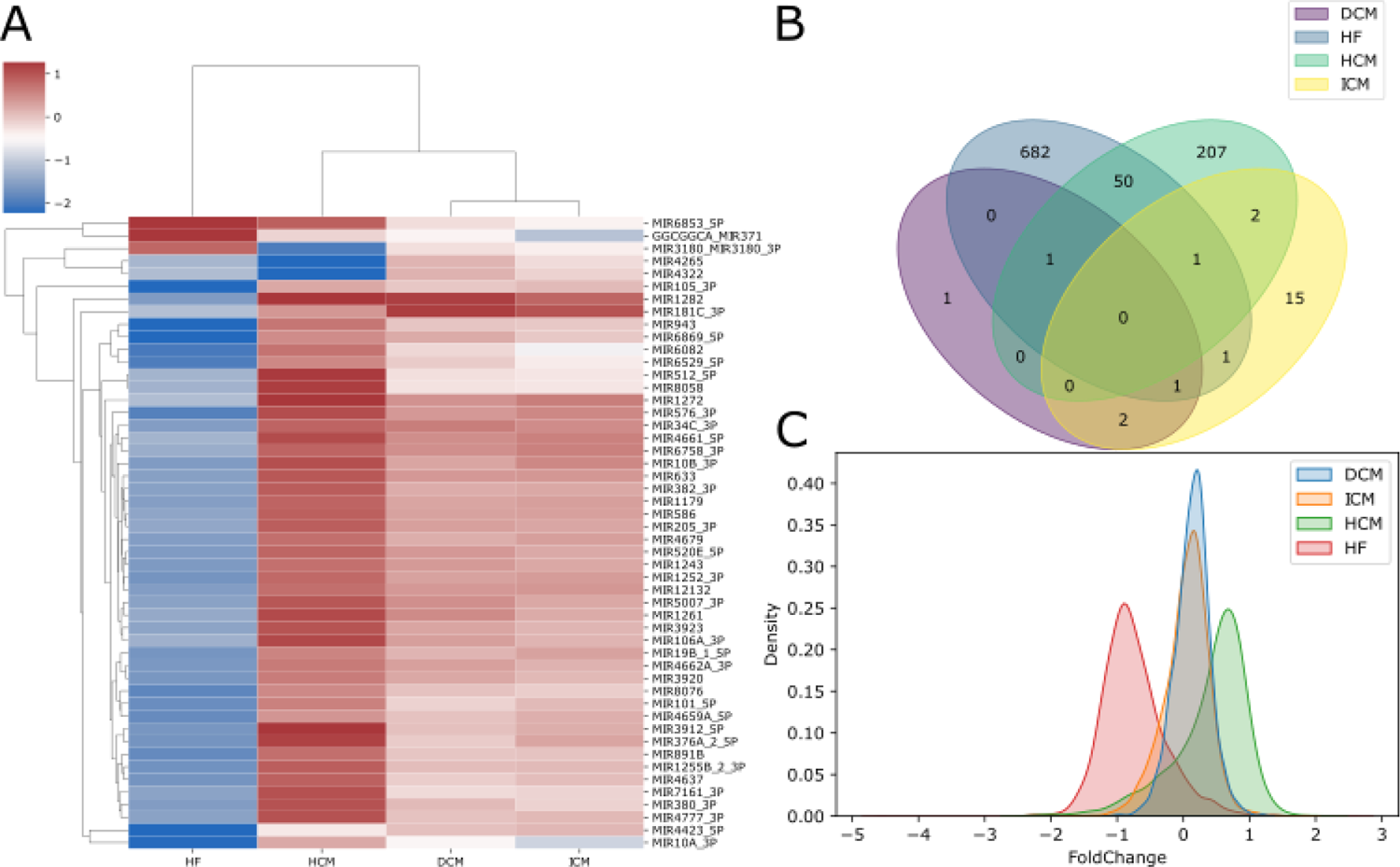
(A) The heatmap shows the pooled effect sizes resulting from the meta-analysis of miRNAs integrated across all studies for each pathology using the random-effects model; (B) The Venn diagram illustrates the intersection of differentially expressed miRNAs across heart failure, dilated cardiomyopathy, ischemic cardiomyopathy, and hypertrophic cardiomyopathy. (C) Distribution of miRNA effect sizes in each cardiac disease.

Conversely, hundreds of differentially expressed miRNAs were identified in HF and HCM. In particular, 736 were found in HF, 728 of which downregulated and 8 upregulated; 261 were found in HCM of which 202 up-regulated and 59 downregulated (Fig. 4c).

Our analysis highlighted various dysregulated miRNAs in HCM, aligning with findings in the current literature, including miR-212, miR-132, miR-22 and miR-199a. Specifically, miR-212 and miR-132 have been identified as both necessary and sufficient for inducing hypertrophic growth in cardiomyocytes. These miRNAs target the expression of the FOXO3 transcription factor, a potent anti-hypertrophic and pro-autophagic factor in cardiomyocytes. The reduction of FOXO3, induced by the increased expression of miR-212/132, leads to the heightened activation of the pro-hypertrophic calcineurin/NFAT signaling pathway, ultimately resulting in the hypertrophy of cardiomyocytes. Conversely, the genetic loss-of-function of miR-212/132, or the use of an antagomir to reduce miR-132, suppresses pressure-overload-induced calcineurin/NFAT signaling, thereby attenuating the development of cardiac hypertrophy ^60^. The increased expression of miR-132, in turn, downregulates autophagy in cardiomyocytes^61^. Another key regulator of autophagy is mTOR. It has been observed that the upregulation of its translational regulator miR-199a alone is sufficient to inhibit cardiomyocyte autophagy and induce cardiac hypertrophy in vivo. These findings reveal a novel role for miR-199a as a crucial regulator of cardiac autophagy, suggesting that targeting miRNAs that regulate autophagy could be a potential therapeutic strategy for treating cardiac diseases ^62^. From our analysis, miR-1282 and miR-7112_5p emerge as the top two upregulated miRNAs in HCM patients when compared to healthy samples. Notably miR-1282 has also been found to be highly upregulated in DCM and it exhibits the opposite behavior in HF, where it is significantly downregulated. Moreover, miR-3912 and miR-3180 emerge as the most upregulated miRNAs in patients affected by ICM and HF. Interestingly, these two microRNAs have been previously associated with acute myocarditis and the growth and metastasis of hepatocellular carcinoma, respectively ^63,64^. Despite their prominent dysregulation, there are currently no studies in the literature linking these miRNAs to ischemic cardiomyopathy or heart failure. This suggests that further studies are necessary to understand their potential involvement in cardiac disease.

### Variations in transcription factor activity profiles

We conducted a differential transcription factor activity analysis for each cardiac disease under study, contrasting affected vs healthy samples, and then compared the results among the different pathologies. Figure 5b shows that only one transcription factor is dysregulated across all four pathologies, emphasizing that the majority of the differences are specific to each pathology. More in detail, in our analysis, Mitogen-Activated Protein Kinase Kinase 1 (MAP2K1) appears to be dysregulated in hypertrophic cardiomyopathy leading to heart failure (Fig 5a). More broadly, the regulation and function of the cardiomyocyte kinome is of great interest, even though few studies exist on the topic. The role of kinases as potential therapeutic targets is an active area of research ^65–67^ and a deeper comprehension of the regulatory networks in which they are involved is needed for understanding the observed cardiac toxicities of some kinase inhibitors.

**Fig. 5.**
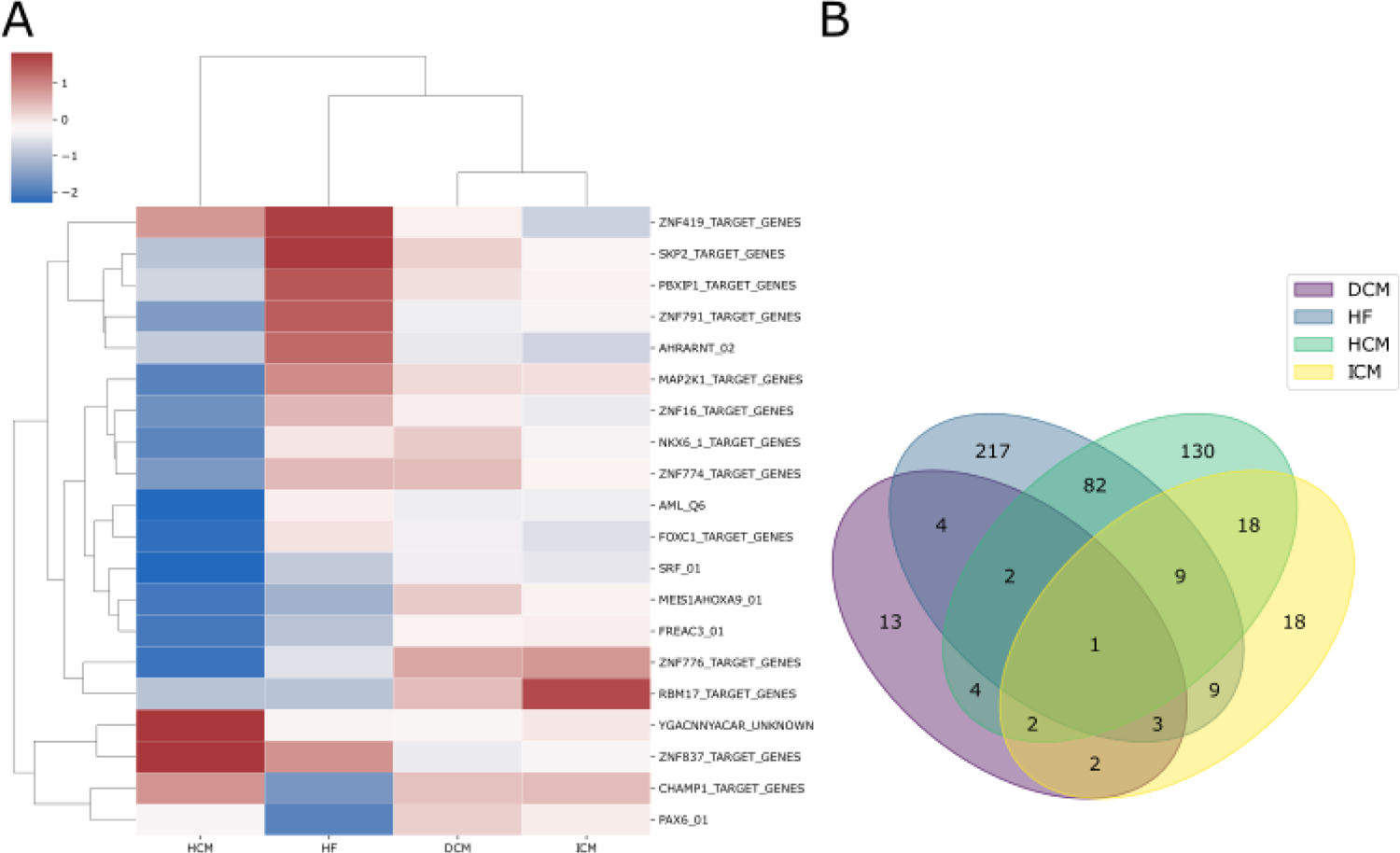
(A) The heatmap shows the pooled effect sizes resulting from the meta-analysis of transcription factor activities computed by GSVA and then integrated across all studies for each pathology using the random-effects model. This analysis further confirms the similarity between DC and ICM, now at the level of transcription factor activity; (B) The Venn diagram shows the intersection of differentially activated TF across heart failure, dilated cardiomyopathy, ischemic cardiomyopathy, and hypertrophic cardiomyopathy.

Although the precise role of MAP2K1 in HF has not been completely described yet, multiple studies reported this kinase to be dysregulated in dilated cardiomyopathy and ischemic heart failure (IHF) ^68,69^. Furthermore, by using bioinformatics and machine learning approaches, Guo and Xu identified MAP2K1 as part of a signature of genes with outstanding diagnostic power, which can discriminate between left ventricular tissue from healthy controls and patients with left heart failure ^69^.

A proposed functional role for MAP2K1 in HF has been suggested in the context of the insulin pathway. There is evidence that the insulin signaling pathway becomes inactive during the onset of heart failure, with MAP2K1 being a pivotal downstream gene within this pathway ^70^. The heart depends on insulin signaling to regulate vital processes such as growth and survival, therefore deficiency in this pathway results in an energy deficit within the heart that accelerates the development of heart failure ^69^.

FOXC1 also emerges as a dysregulated transcription factor from our analysis (Fig 5a). The role of this Transcription Factor in HF possibly relates to the myocardial fibrosis process, defined as the excessive deposition of extracellular matrix in the cardiac interstitium, proliferation of cardiac fibroblasts, tissue repair, and scar formation ^71^. Heart failure is often coupled with cardiac remodeling, a process characterized by various pathological changes, chief amongst which is myocardial fibrosis ^72^.

Zhang et al. (2019) found that, in myocardial ischemia, FOXC1 upregulates the expression of Toll-like receptors ^73^ such as TLR4, a member of the interleukin-1 receptor family ^73^. Therefore, FOXC1 acts as an important regulator of the inflammation process. Activation of TLR4 leads to the progression of cardiac hypertrophy and tissue damage ^74^. In accordance with this evidence, Tao at al. found TLR4 to be a hub gene, capable of discriminating between HF and healthy human cardiomyocytes ^72^. This observation suggests that FOXC1, as a regulator of TLR4, may play a role in the fibrotic processes associated with the development of heart failure.

Atrial fibrillation (AF) is the most common arrhythmia in clinical practice and leads to many serious complications, including heart failure. Multiple studies have investigated the molecular mechanisms involved in AF development and progression, highlighting the role of transcription factors in this process.

PBXIP1 has been shown to be an AF-associated biomarker in human left atrial appendages^75^. In our analysis, we further observed an increased activity of this transcription factor in patients with heart failure compared to healthy individuals and a decreased activity in patients with hypertrophic cardiomyopathy, as reported in Figure 5a. These findings underscore the need for further investigation to elucidate its potential implications in atrial fibrillation and cardiac health.

PAX6 and AHRARNT have emerged as interesting nodes in the regulatory network constructed by searching for interaction between transcription factors and miRNAs targets associated with the development of atrial fibrillation ^76^. More specifically, when comparing AF patients with controls, PAX6 was found to be regulated by the downregulation of miR-223-3p, probably through the modulation of AHRARNT transcription factor activity. Furthermore, PAX6 is known to be associated with the regulation of apoptosis in AF.

Finally, our results include multiple dysregulated ZNF proteins in HF conditions compared to controls (Fig 5a). While certain TFs from this family are already recognized for their role in cardiac remodeling, to the extent that they are employed in the treatment of post-infarction cardiac remodeling ^77^, there is still limited evidence in the literature for the ZNF proteins identified in our analysis. The complete results for this analysis are reported in Supplementary S4.

### Construction of the gene regulatory network in heart diseases

The analyses described in previous paragraphs successfully identified significant differences across multiple cardiac pathologies at different layers of transcriptional and translational regulation, including individual genes, TFs, miRNAs, and biological pathways.

Building upon these findings, we next sought to establish connections across these layers, to paint a comprehensive picture of transcriptional and translational regulation in cardiac disease. This holistic approach is necessary to gain a deeper understanding of the underlying pathological processes, and to advance our knowledge of disease etiology. GRNs have been successfully employed to construct regulatory networks for different pathologies, including Parkinson’s disease ^78,79^ and autoimmune diseases ^80^.

The workflow for constructing a Gene Regulatory Network (GRN) in the context of heart diseases is summarized in Figure 1 eart diseases are marked by substantial differences in gene expression within cardiac tissues. In this context, we established an artificial hierarchical GRN framework to uncover potential regulatory associations among the DEGs. We thus selected biological pathways related to cardiac disease, as identified by our pathway analysis and applied a partial correlation algorithm to identify the key regulators of these pathways. Given the exponential rise in possible regulatory relationships with the increasing number of DEGs, the construction of the GRN was centered on genes associated with pathways identified as dysregulated in the above-mentioned pathway activity profiling analysis. Our analysis thus incorporates pathways such as NOTCH, TGF-β, epithelial-to-mesenchymal transition, and K-Ras pathway, alongside those associated with fatty acid metabolism, oxidative phosphorylation, and adipogenesis. Based on our research, these pathways are central to the dysregulation of cardiac tissue homeostasis, showing distinct alteration patterns within various heart failure sub-pathologies. Notably, the NOTCH, TGF-β and K-Ras pathways are associated with inflammation ^81–83^, making them promising targets for focused research, offering valuable insights into cardiac pathology.

The construction of the GRN for heart diseases started from the DEGs previously identified, in a bottom-up approach. The DEGs were categorized into two groups: (i) regulatory genes, such as those encoding transcription factors (TFs), and (ii) structural genes, including those encoding enzymes. The structural genes were used as the bottom layer during GRN construction. Because genes involved in the same biological process may be regulated by the same TF ^84^, correlation coefficients (CCs) were computed for each gene pair in the bottom layer. Co-expressed gene pairs were considered to be regulated by the same TF (see methods). Subsequently, we computed partial correlation coefficients (PCCs) for each co-expressed genes pair by introducing each TF into the analysis (see methods). Using this approach, we identified the first layer of TFs that may directly regulate the bottom genes. This second layer of TFs was then used as input to another Partial Correlation analysis to identify its potential regulators. The transcription factors comprising the GRN, can thus be classified into three categories: i) those regulating only structural genes; ii) those regulating both structural genes and other TFs; and iii) those regulating only other TFs. Finally, each transcription factor was linked with differentially expressed microRNAs targeting it. The list of genes forming the GRNs of each of the analyzed pathologies is reported in Supplementary 5.

The outermost layer includes TFs that regulate the expression of most of the genes involved in the pathways found dysregulated in each of the individual cardiac pathologies, thus suggesting these TFs as potential pharmacological targets. KLF10 emerges as central TF from our analysis in both HF and HCM (Fig 6-7). Additionally, in both GRNs, KLF10 is associated with miR-130a_5p, which is found to be dysregulated in affected patients. According to current literature, KLF10 is expressed in specific cell types in a wide variety of tissues, and it is known to be involved in repressing cell proliferation and inflammation as well as in the induction of apoptosis ^85^. Furthermore, its involvement in the development of hypertrophic cardiomyopathy in mice has been demonstrated ^86^.

**Fig 6.**
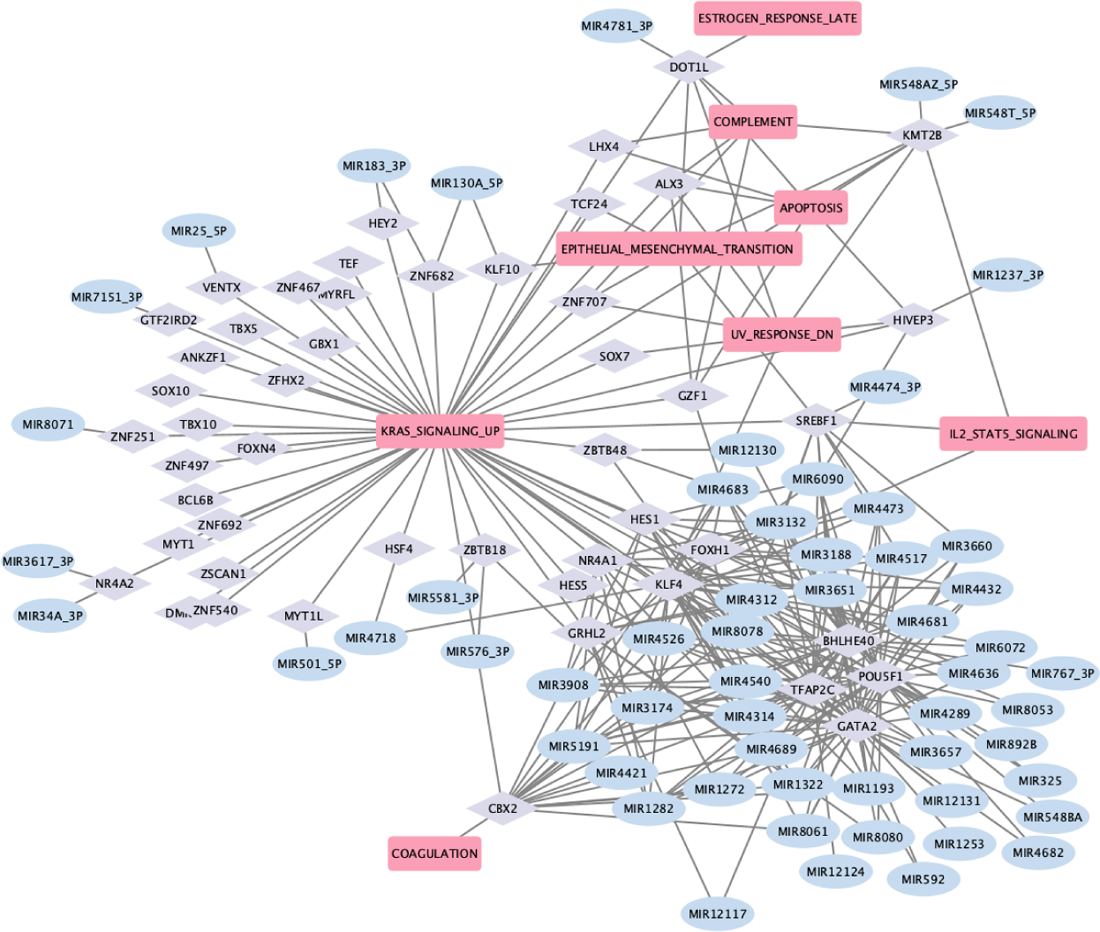
GRN Heart Failure. The rounded pink rectangles depict differentially active pathways in patients with HF; the light violet diamond nodes represent transcription factors involved in the regulation of DEGs; the light blue ellipses represent dysregulated miRNAs that regulate the transcription factors.

**Fig 7.**
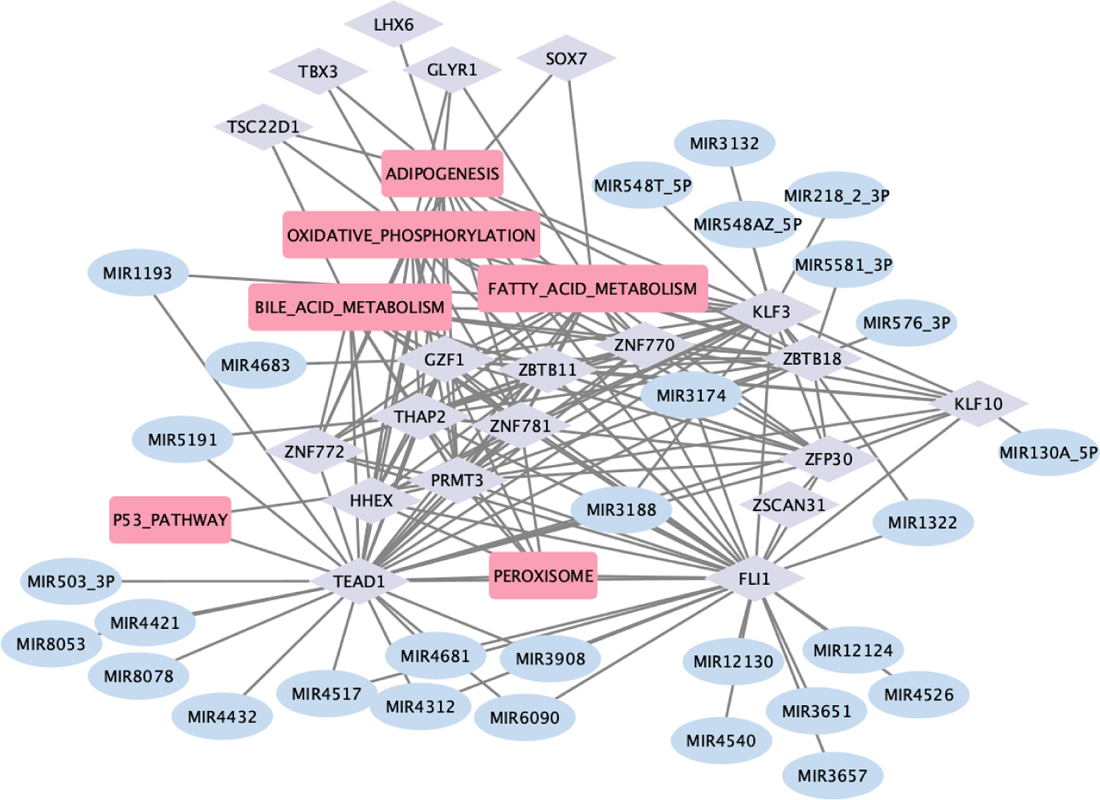
GRN Hypertrophic Cardiomyopathy. The rounded pink rectangles depict differentially active pathways in patients with HCM; the light violet diamond nodes represent transcription factors involved in the regulation of DEGs; the light blue ellipses represent dysregulated miRNAs that regulate the transcription factors.

Our GRN also highlights the importance of TBX3 in ischemic cardiomyopathy (Fig 8). Mutations in this gene have been associated with both congenital and acquired heart diseases ^87,88^. However, no clear association with ischemic cardiomyopathy has been reported.

**Fig 8.**
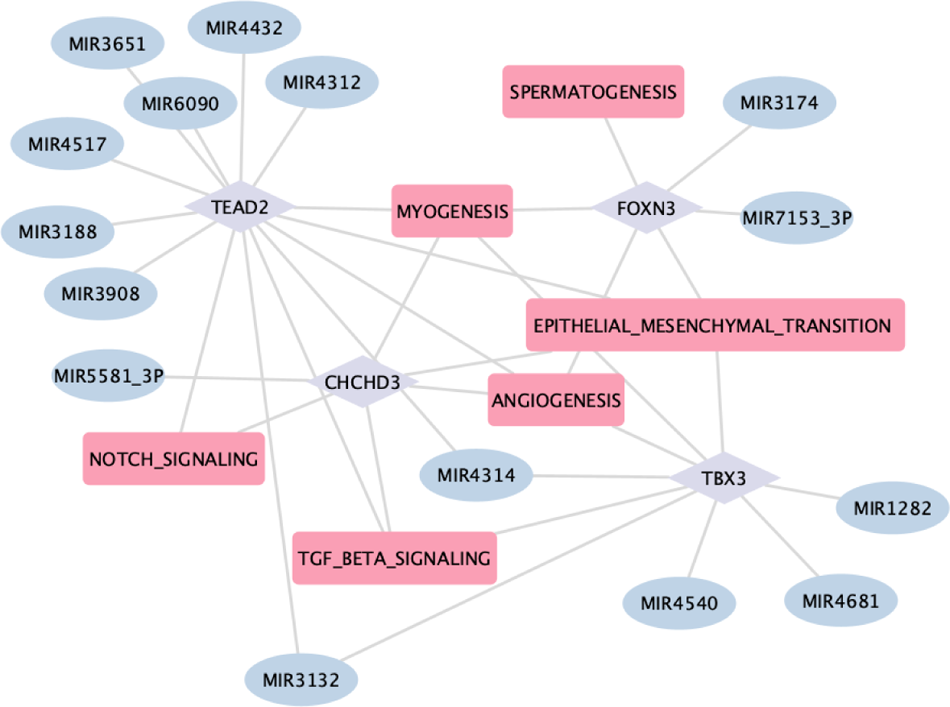
GRN Ischemic Cardiomyopathy. The rounded pink rectangles depict differentially active pathways in patients with ICM; the light violet diamond nodes represent transcription factors involved in the regulation of DEGs; the light blue ellipses represent dysregulated miRNAs that regulate the transcription factors.

Other interesting transcription factors identified in this analysis, associated with both ischemic and dilated cardiomyopathy, are TEAD1 and TEAD2 (Fig 8-9), which are robustly expressed in the embryonic and early postnatal heart and continue to be expressed in the adult heart ^89^. The GRN highlights that TEAD1 is a target of various dysregulated miRNAs in DCM affected-samples, as depicted in Figure 9. Additionally, TEAD1 is a key transcription factor involved in the regulation of several DEGs, which are part of dysregulated pathways in DCM. Despite the decrease of its expression levels with age, TEAD1 maintains a crucial role in regulating cardiac processes, and its dysregulation has been associated with the development of dilated cardiomyopathy in vivo ^90^. The presence of Transcription Factors that have been reported in other publications validates the structure of our GRN, which also includes multiple factors that, to the best of our knowledge, have not been previously implicated in heart disease and warrant further study.

**Fig 9.**
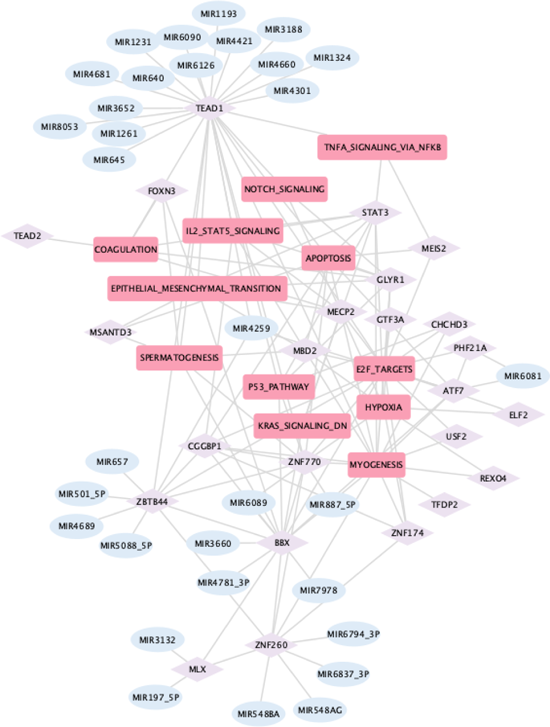
GRN Dilated Cardiomyopathy. The rounded pink rectangles depict differentially active pathways in patients with DCM; the light violet diamond nodes represent transcription factors involved in the regulation of DEGs; the light blue ellipses represent dysregulated miRNAs that regulate the transcription factors.

## Conclusions

In conclusion, our proposed PCC-based algorithm was able to reconstruct a specific Gene Regulatory Network (GRN) that captures transcriptional changes in multiple cardiac pathologies. Notably, several transcription factors highlighted by our approach, such as FOXC1 in heart failure and KLF10 and KLF15 in ischemic cardiomyopathy, are supported by existing literature, validating our approach. The multi-cohort analysis of cardiac pathologies revealed that ischemic and dilated cardiomyopathies exhibit the highest degree of similarity to each other in terms of their patterns of differential gene expression, transcription factor and pathway activity. Moreover, the GRN describes complex regulatory associations among transcription factors (TFs) and between TFs and both miRNAs and structural genes in cardiomyopathies and heart failure. Within this network, dysregulated TFs, such as TEAD1, TEAD2 are implicated in specific pathways linked to cardiac adaptation, including fatty acid metabolism, oxidative stress response, epithelial-to-mesenchymal transition, angiogenesis and coagulation. Our GRNs highlight the importance of TBX3 in ischemic cardiomyopathy and FOXC1 in heart failure, both of which were previously associated with cardiac disorders but not specifically linked with cardiomyopathies or heart failure. The pivotal roles of these TFs in initiating or progressing various cardiac diseases underscore their potential as therapeutic targets. The power of our study is limited by the number of publicly available studies that include both transcriptomic and miRNA data for cardiac pathologies. Moreover, while our meta-analysis framework identifies signals that are consistent across heterogeneous datasets, thus minimizing dataset-specific confounders, it is also possible that smaller effects may be missed due to the heterogeneity of the data. Additionally, the lack of longitudinal data precludes an understanding of the evolution of the disease over time. These limitations notwithstanding, the GRNs assembled in this study delineate key regulatory axes in cardiac pathology and suggest a number of targets for pharmacological intervention that deserve further investigation.

## Materials and methods

### Dataset collection

The datasets related to human heart diseases were collected from the Gene Expression Omnibus (GEO) database. We collected all the six studies in GEO that include both bulk gene expression data and microRNA expression profiles in patients with heart failure, ischemic cardiomyopathy, dilated cardiomyopathy, and hypertrophic cardiomyopathy (GSE55296, GSE116250, GSE133054, GSE135055, GSE48166, GSE55296).

### Gene set variation analysis (GSVA)

The GSVA analysis between patients and healthy controls was performed using the GSVA R package ^91^ by using the Hallmark gene sets ^92^ as the reference gene set and setting the P value to < 0.05 and the t value to >= 1 as the cut-off criteria.

### Meta-analysis

We used the *MetaIntegrator* R package ^93^ to integrate discovery cohorts and identify differentially expressed features between healthy controls and patients with different heart conditions. We first computed effect sizes for each gene and microRNA, as well as for pathways and transcription factors, based on gene set activity in each study. Next, we summarized the effect sizes across all studies for each feature, by weighting the effect size according to the inverse of the variance in that study. Finally, we performed multiple hypothesis correction on the p-values for the summary effect size of each feature by using the Benjamini-Hochberg false discovery rate (FDR). We used the following thresholds in our meta-analysis to select genes in the heart disease MetaSignature: absolute value of effect size > 1, and FDR < 0.05.

### Hierarchical gene regulatory network

We employed a partial correlation coefficient-based algorithm (PCC) to construct the gene regulatory network based on transcription factors. Firstly, we identified co-expressed structural genes using a Pearson correlation coefficient threshold of (r) ≥ 0.7. Subsequently, we tested that the correlation was not influenced by the effect of a third variable. This makes it possible to identify whether the correlation rxy between the variables x and y is caused by a third variable z.The partial correlation rxy,z tells how strongly the variable x correlates with the variable y, if the correlation of both variables with the variable z is factored out 94. A gene pair was then identified as regulated by the same transcription factor if PCC <=0.4. To integrate the results obtained from each individual dataset, we employed a meta-analysis approach. For each pair of genes, we calculated the correlation of their expression in each individual dataset. Subsequently, we integrated the results from the various studies to generate a comprehensive analysis. This method allowed us to consolidate findings across multiple datasets, providing a more robust view of the relationships between multiple genes. For each disease, we created a weighted average co-expression matrix by computing the mean of correlations for each gene pair across all datasets. Each correlation value was assigned a weight based on its corresponding standard error. This approach ensured that more reliable and precise correlations carried more influence in the construction of the co-expression matrix. Given the sample sizes, the standard error was calculated as (1 − *r*^2^) / √*N* − 3, where *r* is the correlation coefficient and *N* is the number of samples in each dataset 95. We applied a similar procedure to calculate the weighted average PCC.

## Article Information Disclosures

None

## Supplementary Material

Supplementary S1

Supplementary S2

Supplementary S3

Supplementary S4

Supplementary S5

## Data Availability

The authors confirm that the data supporting the findings of this study are available within the article [and/or] its supplementary materials.

